# Effect of surgical approach to hip arthroplasty on postoperative pain and mobilization and on efficacy of intrathecal oxytocin for pain

**DOI:** 10.1101/2025.02.22.25322560

**Authors:** John S. Shields, D. Chiamaka Ijebuonwu, Elizabeth G. Korn, Ariel Mueller, Timothy T. Houle, Maxwell K. Langfitt, David C. Pollock, James C. Eisenach, Spinal Oxytocin Hip Surgery Collaborators

**Affiliations:** Department of Orthopaedic Surgery and Rehabilitation, Wake Forest University School of Medicine, Winston-Salem, NC; Department of Anesthesia, Critical Care and Pain Medicine, Massachusetts General Hospital, Harvard Medical School, Boston, MA; Morehouse School of Medicine, Atlanta, GA; Department of Anesthesiology, Wake Forest University School of Medicine, Winston-Salem, NC

**Keywords:** Oxytocin, neuropathic pain, analgesia, spinal injection, hyperalgesia, allodynia

## Abstract

**Objective:** Compare the effect of posterior surgical approach (PA) vs direct anterior approach (DAA) on speed of recovery from pain and dysfunction and on intrathecal oxytocin analgesia after total hip arthroplasty (THA).

**Study design:** Nested cohort within a randomized, controlled, double-blind trial

**Setting:** Hospital

**Subjects:** Individuals aged 31 to 80 years undergoing total hip arthroplasty (THA)

**Methods:** In this secondary analysis of a randomized controlled trial, the association between surgical approach and number of daily steps was assessed, and whether this was modified by receipt of intrathecal oxytocin. Data were collected from accelerometers and daily patient diaries in the first eight weeks postoperatively. Outcomes were analyzed using generalized linear regression models.

**Results:** Ninety patients underwent THA, of which 35 (38.9%) received a PA. Patients were predominantly female (57.8%) with a mean age of 60.6 (standard deviation [SD] 9.3) years. On postoperative day one patients who received a PA with placebo took more steps (mean difference [MD] 53.72, 95% CI: −1717.86, 1825.31) than patients who underwent a DAA. Trajectories were significantly modified by whether they received oxytocin, in which patients who underwent PA with oxytocin took more steps than patients who underwent DAA (p<0.001).

**Discussion:** Further studies are needed to understand mechanisms underlying oxytocin’s interaction with surgical approach and guide considerations for recovery after THA.

## Introduction

Total hip arthroplasty (THA) is one of the most common and successful procedures done in orthopedics with an estimated 262,000 cases per year, which is expected to grow by 176% in 2040 and 659% by 2060. [1] The most common surgical approaches utilized are the direct anterior approach (DAA) which utilizes an internervous plane, and the posterolateral approach (PA) utilizing an intramuscular plane. The posterolateral approach is most commonly performed, but the DAA has gained in popularity given multiple publications citing improved advantages of less pain, faster return to function, and lower dislocation risk.[2-5] Proponents of the PA reference studies that show no difference between the two approaches.[6-8] Regardless of approach, surgeons continue to search for avenues to improve pain and function after THA as volumes shift to outpatient surgery.

Research on recovery after THA increasingly utilizes patient centered functional and activity outcomes and remote monitoring devices, which are well accepted and may in themselves be therapeutic [9]. Despite the presence of daily or near continuous measurement frequency, however, most reported outcome measures utilize values at only a few times (e.g., weekly or monthly) [10]. In contrast, we recently modeled trajectories of daily measures of pain, opioid use and physical dysfunction for two months after THA in an interventional trial of spinal oxytocin [11]. The primary goal of this pre-specified secondary analysis of that trial is to examine the influence of surgical approach on these individual, data-rich, patient-centered outcomes. It was hypothesized that addition of surgical approach to the model would significantly improve fit and that, compared to PA, patients receiving DAA would have a more rapid and extensive increase in their daily step counts over the eight-week period after THA. It was further hypothesized that parallel benefits of DAA over PA would be observed in secondary outcomes of pain scores, opioid use, self-reported disability, duration and time to ambulation with physical therapy, and hospital length of stay.

## Methods

This study was a nested cohort study within a randomized controlled trial. The parent trial was a prospective, parallel arm, randomized (1:1), blinded, controlled superiority trial comparing oxytocin versus placebo. Surgeries performed in the parent trial took place at two sites within the Atrium Health Wake Forest Baptist Health System in North Carolina between January 2017 and August 2020 [11]. Complete details of the protocol, inclusion criteria of participants, and randomization are available in both the primary publication [11] and at clinicaltrials.gov (NCT03011307). All subjects provided written informed consent for the primary trial and subsequent use of data for analysis. Another pre-specified secondary analysis from this dataset, which examined acute hemodynamic effects of spinal oxytocin, has been previously published [12].

The exposure of interest for the present analysis was surgical approach – defined as either DAA or PA. The sample size was fixed such that all patients included in the primary analysis (defined as all randomized patients who received the study drug) were included in this secondary analysis.

### Outcomes

The primary outcome of this secondary analysis was the number of daily steps taken after surgery. To assess this, patients wore an accelerometer to record daily step counts for eight weeks postoperatively. Baseline step counts were also recorded via an accelerometer for up to two weeks prior to surgery.

Secondary outcomes included worst daily postoperative pain, opioid self-administration, self-reported disability, length of hospital stay, and time to first physical therapy mobilization and distance of ambulation. Postoperative pain was assessed via an electronic diary, in which patients were asked to report the worst pain intensity in the previous 24 hours on a scale from 0 (no pain) to 10 (worst imaginable pain). Postoperative opioid administration was self-reported via daily diary entries and reported as present or absent each day. Self-reported disability was measured weekly via electronic survey using the World Health Organization Disability Assessment Schedule (WHODAS) 2.0 [13]. Time to first physical therapy mobilization was calculated as the time from end of surgery to their first steps at the first physical therapy appointment as recorded in the medical record. Distance of ambulation was defined as the distance (in feet) the patient took at the first physical therapy appointment. In circumstances where distance was inadvertently recorded in steps, two feet was imputed for each recorded step. Length of hospital stay was reported in days and defined as the date of discharge minus date of surgery.

### Statistical Analysis

Descriptive statistics of the data are presented as mean ± standard deviation, median (interquartile range [IQR]) or frequency counts and percentages depending on variable type and distribution. Standardized mean differences (SMD) were reported to demonstrate differences between those receiving a DAA and PA. Superiority testing was used to examine differences between the two surgical approaches for the primary outcome and all secondary outcomes. Hypothesis testing was two-tailed with statistical significance interpreted at p < 0.05. Hierarchical linear mixed effect models, with distributions appropriate to the variable under study, were utilized for analysis including a random intercept for each subject. All models were adjusted for prognostic covariates including age, sex, study site where the procedure was performed, postoperative day (using a natural log transformation) and the interaction with prognostic covariates. This model framework allows for estimation of the intercept (the day after postoperative discharge) and the changes in the trajectory over time (slope) in each variable. Additionally, while the attending surgeon plays a large role in determining the surgical approach, it was not included in the models because surgical approach co-varied strongly with surgeon.

To evaluate the primary outcome, hierarchical linear models were constructed and evaluated in the following ways. First, the number of daily steps was regressed onto prognostic covariates and randomization group (i.e., oxytocin or placebo), and the interaction with the natural log of the postoperative day (Model 1). Then, surgical approach was incorporated as a fixed effect, including its interaction with time (Model 2). Lastly, in Model 3, an interaction term was included for surgical approach and oxytocin (and their interaction with time) in order to evaluate whether receipt of oxytocin modified the relationship between surgical approach and outcome. Likelihood ratio tests were used to evaluate model fit for the primary inference, which evaluated model fit between Model 1 and Model 2. Presence of the interaction between approach and oxytocin was further evaluated by comparing Models 2 and 3. Results are presented as a mean difference (MD) and its associated 95% confidence interval (CI). A sensitivity analysis was performed of the best fitting model, including adjustment for the average number of steps taken at baseline in the two weeks prior to surgery.

Secondary outcome measures were evaluated using the same model framework and covariates, including distributions and link functions appropriate for the outcome. This included a gaussian distribution and identity link for steps, pain scores and self-reported disability scores, and a binomial distribution and logit link for opioid administration. Results of the opioid administration model are presented as odds ratios (OR) and 95% CIs. In addition to prognostic covariates, self-reported disability models included an *a priori* adjustment for baseline preoperative disability score. Duration of ambulation was evaluated with linear regression, conditional on randomization strata (oxytocin or placebo) and prognostic covariates, as well as their interaction with surgical approach. Time to discharge and ambulation were evaluated as time-to-event data using Cox Proportional Hazards regression, with results reported as hazards ratios (HR) and 95% CIs. No imputation was performed for missing data. All analyses were performed using R4.2.3 (R Foundation for Statistical Computing, Vienna, Austria) and RStudio 2023.03.0 +386 (RStudio, Inc).

## Results

### Study Participants

Overall, 90 patients underwent a THA, of whom 55 (61%) received a DAA and 35 (39%) received a PA. Meaningful differences in surgeon preferences were observed as the attending surgeon plays a large role in determining the surgical approach taken. Patient characteristics were otherwise well balanced between groups, with the exception of some differences in race, hospital site, and location of pain other than the hip, with patients undergoing PA more likely to be Black, have surgery at the main campus, and have back or spine pain (Table 1).

**Table 1.**
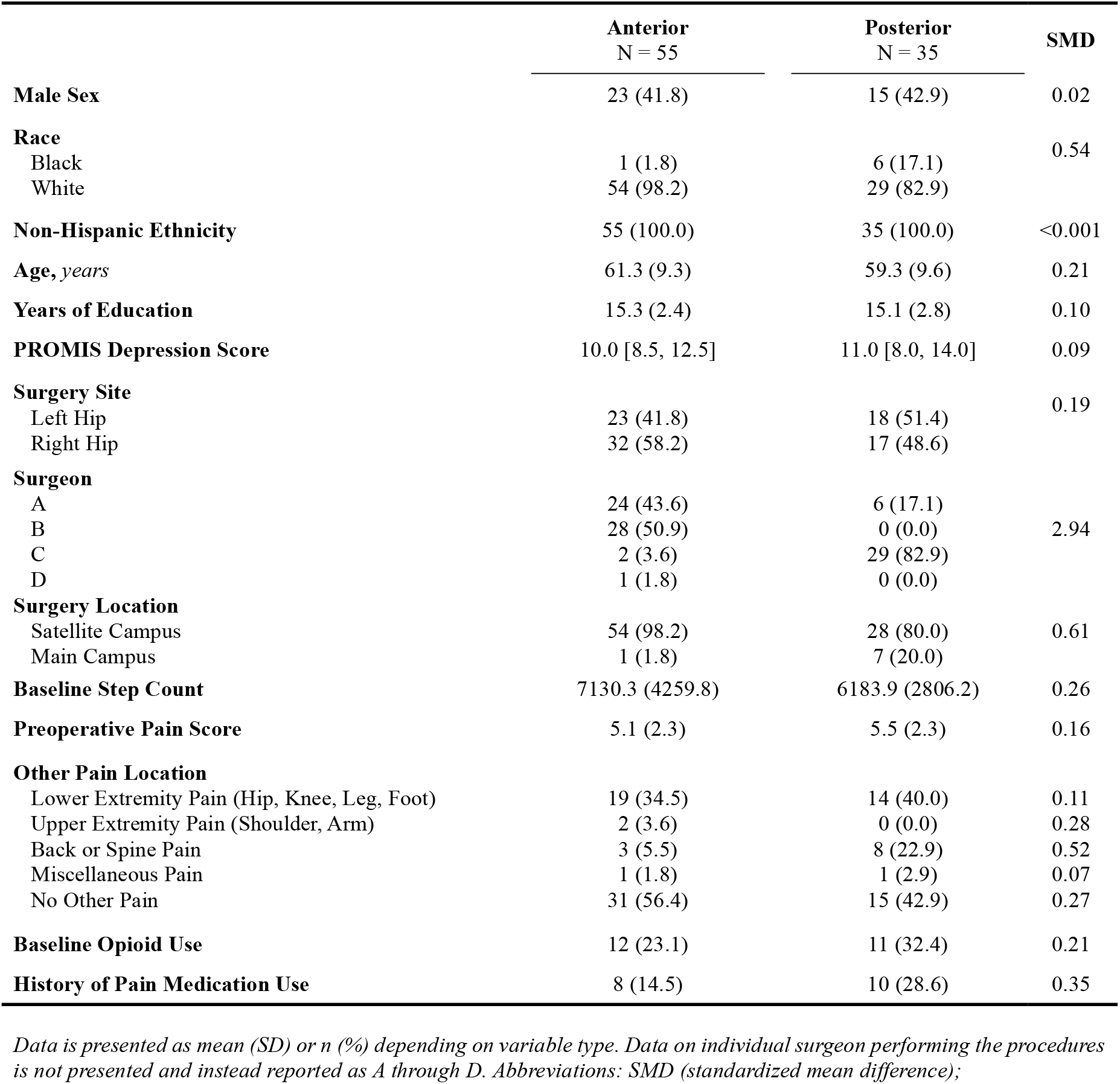
Patient and Surgical Characteristics Stratified by Surgical Approach.

### Primary Analysis: Number of Steps Postoperatively

Of the 90 participants, 85 had step counts available postoperatively and were thus included in the primary analysis. Step counts were collected from postoperative day 1 through day 56 for all 85 participants. Excluding 454 days of missing step data (9.5%), a total of 4,306 days of step count data were included in the analysis of postoperative step counts.

Results from the ANOVA likelihood ratio test in the primary analysis did not suggest that surgical approach impacts the trajectory of recovery after surgery (X^2^ = 0.02, df = 2, p = 0.99). When considering Models 2 and 3 however, this trajectory was significantly modified by receipt of oxytocin (X^2^ = 21.23, df = 2, p < 0.001). As such, patients who underwent PA with oxytocin took more steps than patients who underwent DAA with oxytocin over the eight-week period whereas those receiving placebo took fewer steps if they underwent a PA (Figure 1; Table 2; p < 0.001). For example, using conditional predictions from the model, a patient who underwent PA with oxytocin took 5495 steps on postoperative day 28, while a patient who underwent DAA with oxytocin took 4537 steps. Alternatively, a patient who underwent PA with placebo took only 4035 steps on postoperative day 28, while a patient who underwent DAA with placebo took 5164 steps. These results were conserved in a post hoc sensitivity analysis that adjusted for average baseline step count in the two weeks prior to surgery (Supplementary Table 1).

**Table 2.**
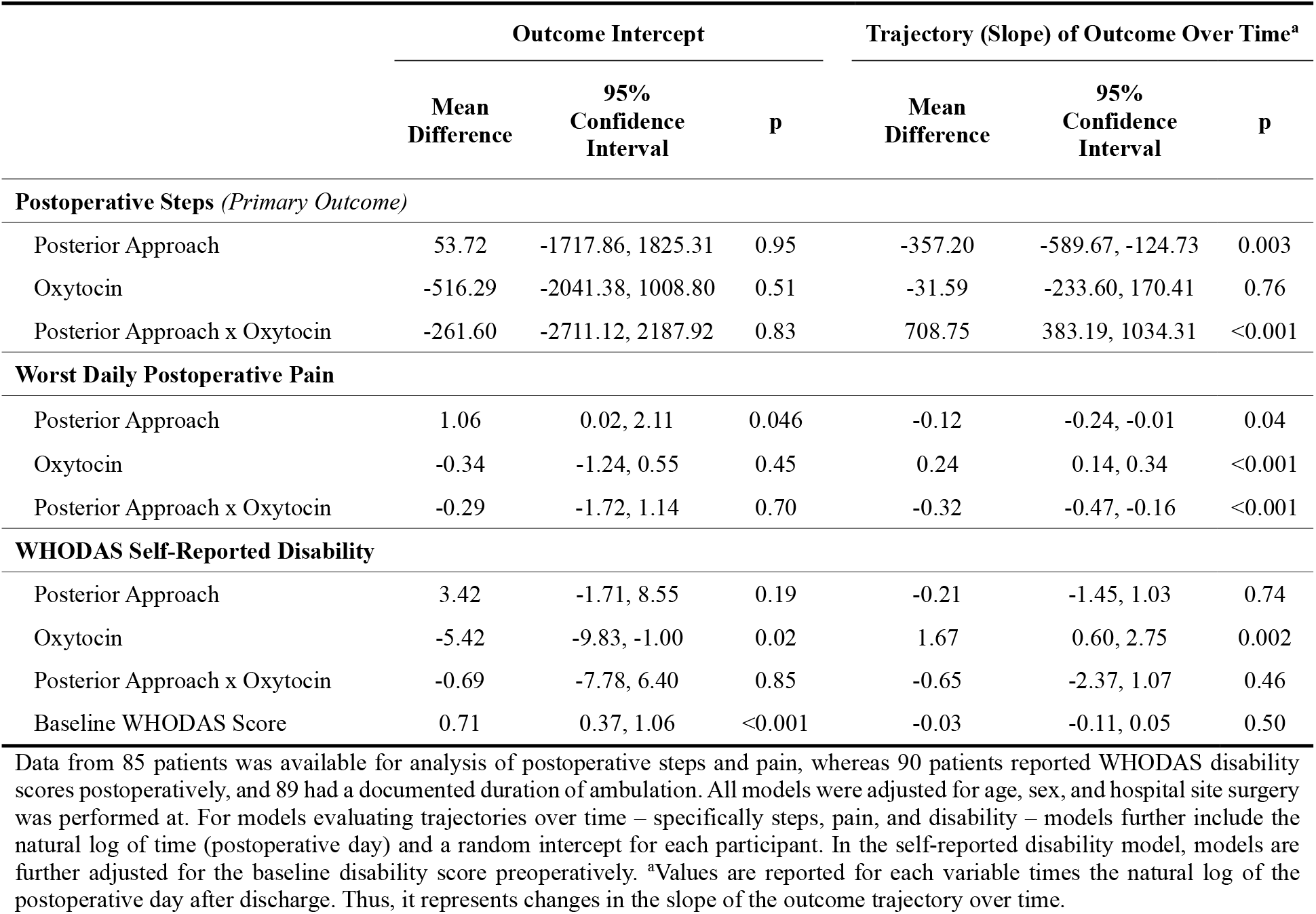
Models for Continuously Scaled Primary and Secondary Outcomes.

|  | Outcome Intercept |  |  | Trajectory (Slope) of Outcome Over Time<sup>a</sup> |  |  |
| --- | --- | --- | --- | --- | --- | --- |
|  | Mean Difference | 95% Confidence Interval | p | Mean Difference | 95% Confidence Interval | p |
| **Postoperative Steps (Primary Outcome)** |  |  |  |  |  |  |
| Posterior Approach | 53.72 | -1717.86, 1825.31 | 0.95 | -357.20 | -589.67, -124.73 | 0.003 |
| Oxytocin | -516.29 | -2041.38, 1008.80 | 0.51 | -31.59 | -233.60, 170.41 | 0.76 |
| Posterior Approach x Oxytocin | -261.60 | -2711.12, 2187.92 | 0.83 | 708.75 | 383.19, 1034.31 | <0.001 |
| **Worst Daily Postoperative Pain** |  |  |  |  |  |  |
| Posterior Approach | 1.06 | 0.02, 2.11 | 0.046 | -0.12 | -0.24, -0.01 | 0.04 |
| Oxytocin | -0.34 | -1.24, 0.55 | 0.45 | 0.24 | 0.14, 0.34 | <0.001 |
| Posterior Approach x Oxytocin | -0.29 | -1.72, 1.14 | 0.70 | -0.32 | -0.47, -0.16 | <0.001 |
| **WHODAS Self-Reported Disability** |  |  |  |  |  |  |
| Posterior Approach | 3.42 | -1.71, 8.55 | 0.19 | -0.21 | -1.45, 1.03 | 0.74 |
| Oxytocin | -5.42 | -9.83, -1.00 | 0.02 | 1.67 | 0.60, 2.75 | 0.002 |
| Posterior Approach x Oxytocin | -0.69 | -7.78, 6.40 | 0.85 | -0.65 | -2.37, 1.07 | 0.46 |
| Baseline WHODAS Score | 0.71 | 0.37, 1.06 | <0.001 | -0.03 | -0.11, 0.05 | 0.50 |

Data from 85 patients was available for analysis of postoperative steps and pain, whereas 90 patients reported WHODAS disability scores postoperatively, and 89 had a documented duration of ambulation. All models were adjusted for age, sex, and hospital site surgery was performed at. For models evaluating trajectories over time – specifically steps, pain, and disability – models further include the natural log of time (postoperative day) and a random intercept for each participant. In the self-reported disability model, models are further adjusted for the baseline disability score preoperatively. <sup>a</sup>Values are reported for each variable times the natural log of the postoperative day after discharge. Thus, it represents changes in the slope of the outcome trajectory over time.

**Figure 1.**
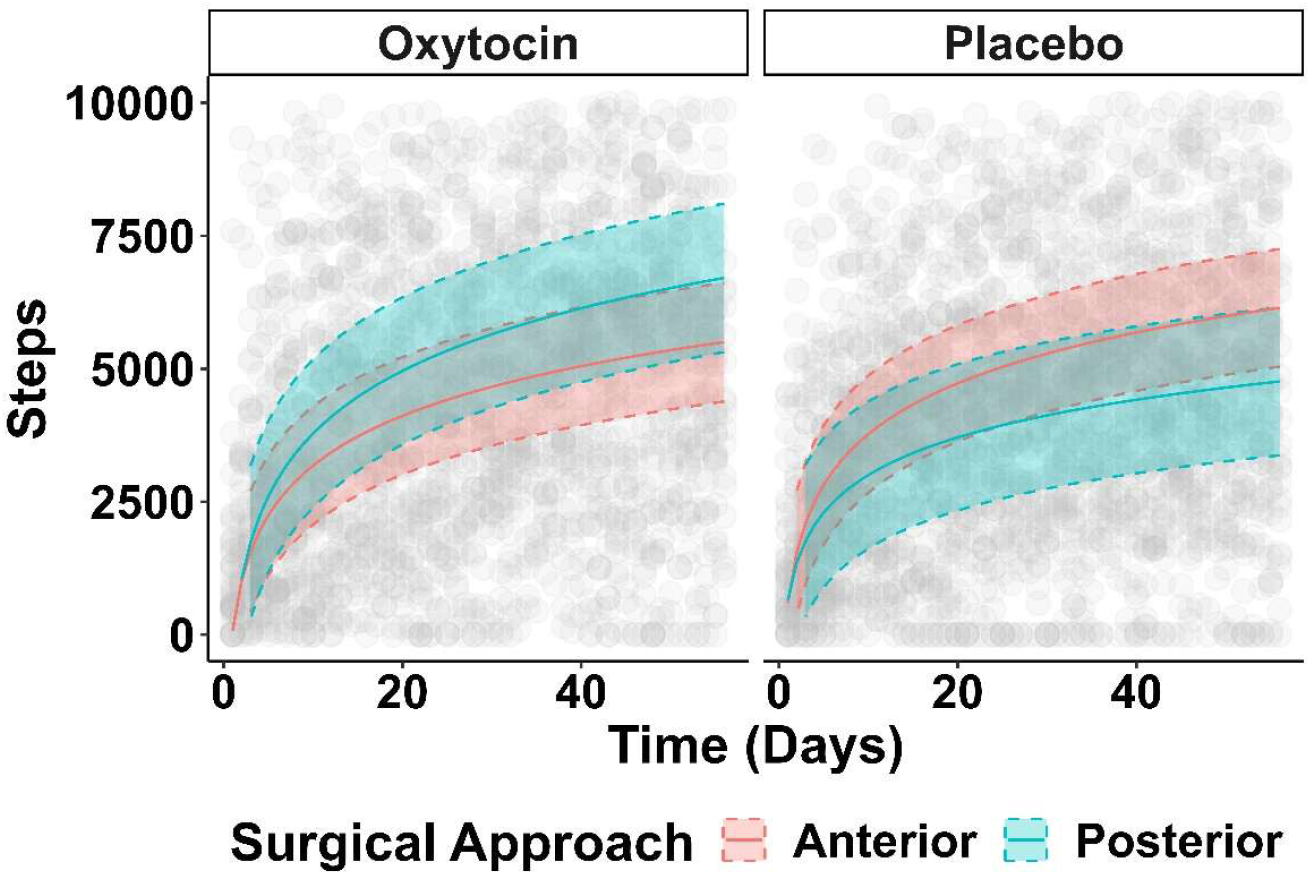
Trajectory of Steps Postoperatively. The number of steps taken in the first eight weeks postoperatively is reported stratified by surgical approach. Patients who received a direct anterior approach are reported in red, whereas those receiving the posterior approach are reported in blue. Given the observed interaction with oxytocin use, this is presented separately for those who received oxytocin (left panel) as compared to those who received placebo (right panel). Step counts for individual patients are reported in grey.

### Postoperative Pain and Opioid Administration

In separate models for pain and opioid use, the same model framework was used as outlined above. Likelihood ratio tests for each model demonstrated that the trajectory of pain and of opioid use was associated with surgical approach (X^2^ = 45.10, df = 2, p < 0.001 and X^2^ = 15.13, df = 2, p < 0.001, respectively), however this was further modified by receipt of oxytocin (X^2^ = 18.99, df = 2, p < 0.001 and X^2^ = 14.48, df = 2, p < 0.001, respectively).

On the day after hospital discharge, patients who received a PA in the placebo group reported greater pain intensity (MD 1.06 points, 95% CI: 0.02, 2.11, p = 0.046; Table 2) but had no difference in the odds of taking opioids at home compared to those who received a DAA (OR 0.03, 95% CI: 0.00, 2.48; p = 0.12; Table 3). When considering the trajectory over time however, patients who underwent a PA with oxytocin reported less pain than those who underwent a DAA with oxytocin whereas those who underwent the PA with placebo reported more pain over the eight-week period (Figure 2A). In addition, patients who received the PA had a significantly increased probability of using opioids irrespective of whether they received oxytocin or not after one week. However, there was significantly more variability in opioid use among those receiving placebo as compared to oxytocin, with more patients in the placebo group who received the PA taking opioids longer (Figure 2B).

**Table 3.**
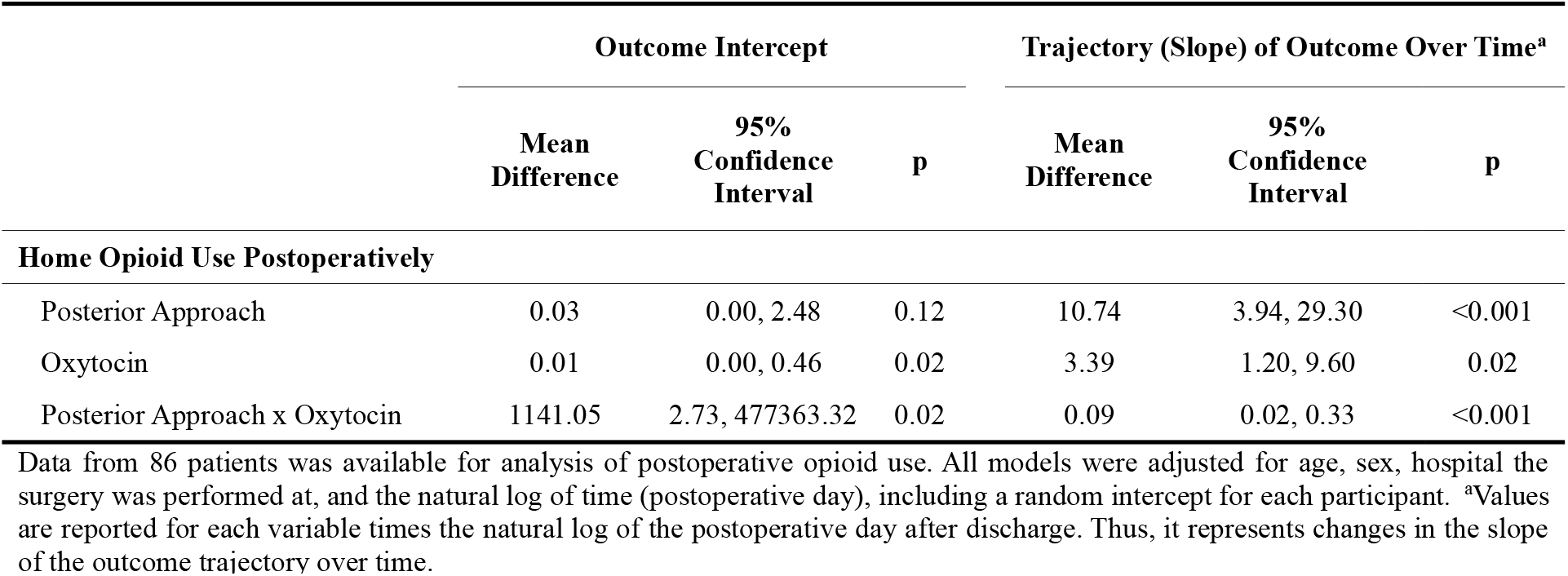
Model for Home Opioid Use.

|  | Outcome Intercept |  |  | Trajectory (Slope) of Outcome Over Time<sup>a</sup> |  |  |
| --- | --- | --- | --- | --- | --- | --- |
|  | Mean Difference | 95% Confidence Interval | p | Mean Difference | 95% Confidence Interval | p |
| **Home Opioid Use Postoperatively** |  |  |  |  |  |  |
| Posterior Approach | 0.03 | 0.00, 2.48 | 0.12 | 10.74 | 3.94, 29.30 | <0.001 |
| Oxytocin | 0.01 | 0.00, 0.46 | 0.02 | 3.39 | 1.20, 9.60 | 0.02 |
| Posterior Approach x Oxytocin | 1141.05 | 2.73, 477363.32 | 0.02 | 0.09 | 0.02, 0.33 | <0.001 |

Data from 86 patients was available for analysis of postoperative opioid use. All models were adjusted for age, sex, hospital the surgery was performed at, and the natural log of time (postoperative day), including a random intercept for each participant. <sup>a</sup>Values are reported for each variable times the natural log of the postoperative day after discharge. Thus, it represents changes in the slope of the outcome trajectory over time.

**Figure 2.**
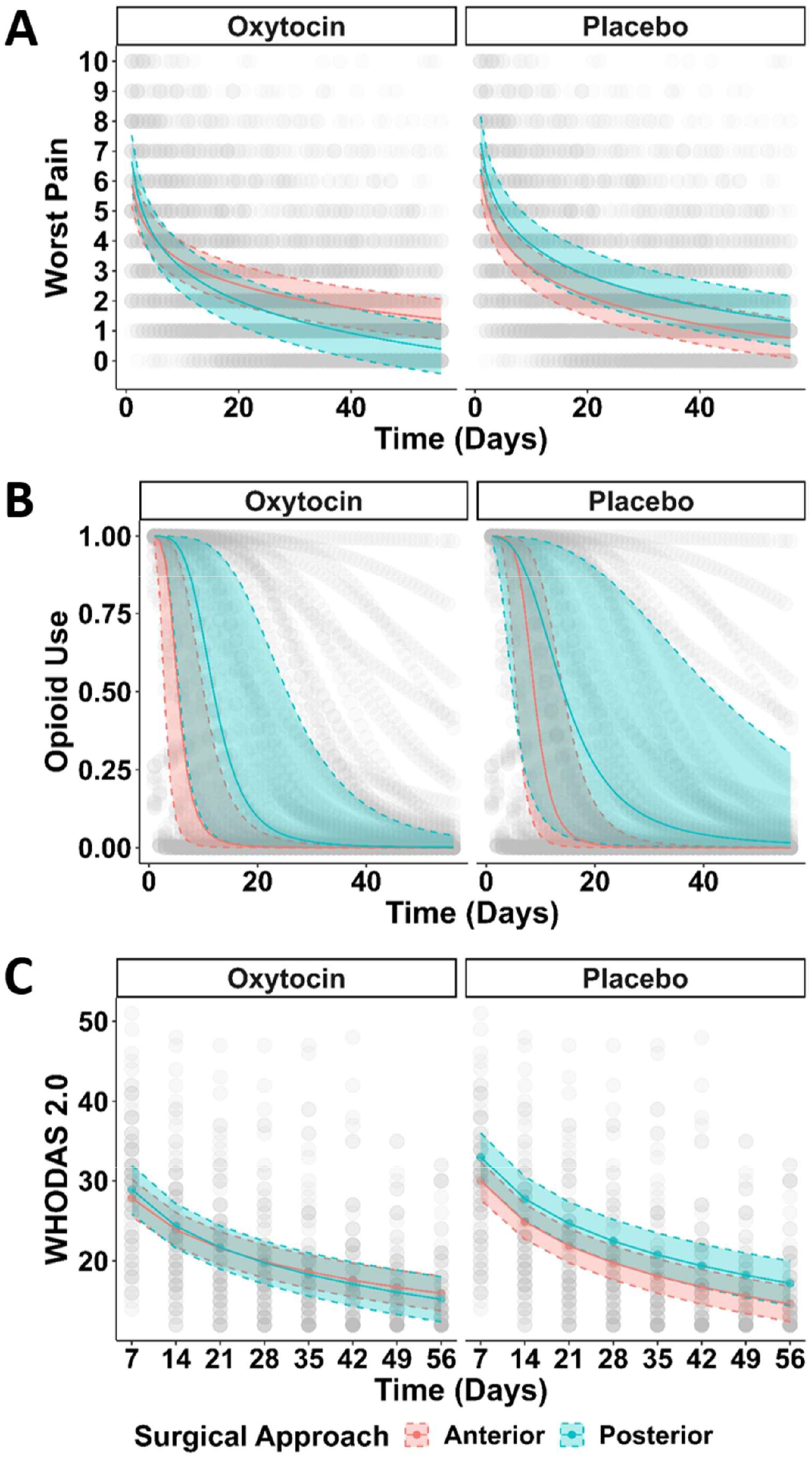
Trajectory of Secondary Outcomes. The worst pain score (Panel A), use of opioids (Panel B) and self-reported disability (Panel C) in the first eight weeks postoperatively is reported stratified by surgical approach. Patients who received a direct anterior approach are reported in red, whereas those receiving the posterior approach are reported in blue. Given the observed interaction with oxytocin use, this is presented separately for those who received oxytocin as compared to those who received placebo. Variability in individual patient responses for each outcome are reported in grey.

### Self-Reported Disability

Results of the likelihood ratio test for self-reported disability suggested that a model incorporating a modest interaction with oxytocin is not significantly different from a model without this interaction (X^2^ = 2.35, df = 2, p = 0.31). In this model, no significant difference was observed in self-reported disability between surgical approach strategies on the day after discharge (Table 2), nor were there meaningful differences in disability scores over time for different surgical approaches.

### Length of Hospital Stay

The median time to discharge was 1 [IQR 1, 1] day in all patients with data available (n = 88). No evidence of an interaction was observed between surgical approach and oxytocin use (X^2^ = 0.50, df = 1, p = 0.48); thus, this was not included in the final model. No significant difference in time to discharge was observed between those who received a PA as compared to patients receiving a DAA (HR 0.71, 95% CI: 0.44, 1.14; p = 0.16; Table 4).

**Table 4.** Models for Secondary Time to Event Outcomes.

|  | Hazard Ratio | 95% Confidence Interval | p |
| --- | --- | --- | --- |
| **Length of Hospital Stay, Days** |  |  |  |
| Posterior Approach | 0.71 | 0.44, 1.14 | 0.16 |
| Oxytocin | 1.22 | 0.79, 1.88 | 0.37 |
| **Time to First Physical Therapy Appointment, Days** |  |  |  |
| Posterior Approach | 0.69 | 0.43, 1.11 | 0.13 |
| Oxytocin | 1.20 | 0.78, 1.84 | 0.42 |

Data from 88 patients was available for analysis of length of stay. All models were adjusted for age, sex, and the hospital in which the surgery was performed. Given the non-significant interactions observed between approach and oxytocin use in the length of stay model ( $X^2=0.50$ , $df=1$ , $p = 0.48$ ) and time to first physical therapy appointment ( $X^2=1.68$ , $df=1$ , $p = 0.20$ ), this was removed from the final models.

### Time to First Physical Therapy and Duration of Ambulation

No evidence of an interaction was observed between surgical approach and oxytocin use for either time to first ambulation with physical therapy (X^2^ = 1.68, df = 1, p = 0.20) or distance of first ambulation (X^2^ = 0.59, df=1, p = 0.46). Additionally, no statistically significant difference was observed for patients who received the PA as compared to the DAA in time to first physical therapy appointment (HR 0.69, 95% CI: 0.43, 1.11; p = 0.13; Table 4) or distance of ambulation at that visit. Patients who received a PA, however, reported a trend towards an increase in the distance of ambulation during the physical therapy visit (MD −48.10 feet, 95% CI: −104.00, 7.80; p = 0.09).

## Discussion

In this study, surgical approach during THA was associated with differences in daily step counts over eight weeks following surgery, although not with differences in time to first physical therapy or distance walked in the acute postoperative period in hospital. Interestingly, whether a patient received oxytocin significantly modified the trajectory of steps over time. Specifically, patients who received oxytocin took more steps if they received the PA, whereas in the context of placebo, patients who received the PA took less steps than those who underwent a DAA. Similar interactions with oxytocin and surgical approach were observed for other secondary outcomes including pain and opioid administration, however no effect modification was observed for self-reported disability, length of hospital stay, distance of or time to ambulation at the first physical therapy appointment.

While THA has been shown to be one of the most successful and cost-effective treatments for hip arthritis, there has been substantial debate over the preferred surgical approach [14]. A previous meta-analysis did not demonstrate any clear superiority of one approach over the other, however four of the included studies highlighted early improvement in pain and function with DAA, with many citing decreased length of stay and dislocation risk [14]. Similarly, in a study of 182 patients, more rapid recovery for hip function and gait was observed after DAA as compared to PA [15]. While Martin et al. also found earlier discharges and mobilization in the DAA group in their retrospective study, a DAA was also noted to have an increased rate of lateral femoral cutaneous neuropraxis (17%) and fracture (2%) [16]. DAA has also been shown to rely less on the use of assistive devices and opioids, earlier discharges, and lower pain scores, but no functional or measurable differences at six weeks as compared to PA [17-19]. Consistent with this previous literature, our study demonstrated improved step counts and pain, and results in a lower use of opioid in the DAA group as compared to the PA group for patients who received placebo.

Oxytocin, a neuropeptide synthesized by neurons in the hypothalamus with projections to the pituitary and to many sites in the central nervous system, produces analgesia in animals primarily by an action in the spinal cord [20]. Although we failed to observe an effect of spinal oxytocin just prior to THA on recovery trajectory in pain, it did speed recovery in activity, reduced disability, and shortened the time of opioid treatment [11]. Oxytocin and placebo groups were well matched for surgical approach in that trial, but we did not test whether efficacy of oxytocin differed according to surgical approach. In this secondary analysis, the interaction between surgical approach and oxytocin was quite striking, yet the mechanism behind this association is not known. It is possible that the effect of surgical approach on efficacy of intrathecal oxytocin could reflect differences in oxytocin disposition after injection. In the parent trial oxytocin was formulated in preservative free saline, which is hypobaric compared with cerebrospinal fluid. As a result, patient position, lateral with the operated hip up in the posterior approach compared to supine in the anterior approach, could conceivably enhance oxytocin distribution and increase its concentration on the operated side, given the hypobaricity of the injectate. Despite this possibility, the effect of position on drug disposition of hypobaric local anesthetics is subtle, and the small difference in oxytocin concentration or spread from positioning could explain the large difference in oxytocin efficacy only if the concentration-effect curve was steep and positioned very close to the concentration of that following acute dilution in lumbosacral cerebrospinal fluid. In addition, injection of local anesthetic very shortly after oxytocin would enhance mixing and render the effects of position smaller. Taken together, although it could be speculated that differences in oxytocin disposition may explain the association we observed, these understandings are quite primitive and future studies would be needed to confirm this mechanistic hypothesis.

This study has limitations. Given the retrospective cohort design nested within the context of a parent randomized controlled trial, it is not possible to rule out the possibility of residual confounding for this secondary analysis. Additionally limiting this study was the small sample size from a single healthcare system, which may affect the generalizability to other surgical settings. In addition, there may be other institutional characteristics that were not measured that could affect the generalizability or interpretation for several of the secondary outcomes we observed. For example, timing of physical therapy or hospital discharge is often multifactorial and may reflect standards of care or other resource considerations that may vary by site. This study did not explicitly evaluate other measures of surgical outcome, nor could it account for surgical provider (because of inadvertently controlling for the exposure it sought to evaluate), therefore it is unclear what role this would play in the observed associations.

Results of this study suggest that surgical approach is associated with differences in number of steps postoperatively and may be associated with pain and opioid use after THA. Importantly, the effect of oxytocin modifies these associations and should be considered when selecting an optimal approach to enhance patient recovery. Future research is needed to truly elucidate these mechanisms and replicate these findings in a larger study population that considers both oxytocin use and surgical approach simultaneously in a robust research design.

## Data Availability

All data produced in the present study are available upon reasonable request to the authors

## Acknowledgements

Supported in part by National Institutes of Health grant R37 GM48085 to JCE.

## Supplementary Appendix 1: Spinal Oxytocin Hip Surgery Collaborators

Regina S. Curry, RN

Sean W. Dobson, MD

Christopher J. Edwards, MD

Daryl S. Henshaw, MD

J. Douglas Jaffe, MD

James D. Turner, MD

Robert S. Weller, MD

All from Department of Anesthesiology, Wake Forest University School of Medicine, Winston-Salem, NC

**Supplementary Table 1.** Post Hoc Sensitivity Analysis.

|  | Outcome Intercept |  |  | Trajectory (Slope) of Outcome Over Time<sup>a</sup> |  |  |
| --- | --- | --- | --- | --- | --- | --- |
|  | Mean Difference | 95% Confidence Interval | p | Mean Difference | 95% Confidence Interval | p |
| **Postoperative Steps** |  |  |  |  |  |  |
| Posterior Approach | 549.18 | -808.35, 1906.71 | 0.43 | -318.32 | -548.98, -87.66 | **0.007** |
| Oxytocin | -98.50 | -1284.66, 1087.66 | 0.87 | 70.19 | -133.56, 273.94 | 0.50 |
| Posterior Approach x Oxytocin | -706.03 | -2607.45, 1195.38 | 0.47 | 649.24 | 323.73, 974.75 | **<0.001** |
| Baseline Steps | -0.07 | -0.20, 0.07 | 0.35 | 0.17 | 0.15, 0.20 | **<0.001** |

Data from 85 patients was available for analysis of postoperative steps. Model was adjusted for age, sex, hospital site surgery was performed at, and mean baseline steps preoperatively. For evaluating trajectory over time, model was further included natural log of time (postoperative day) and a random intercept for each participant. <sup>a</sup>Values are reported for each variable times the natural log of the postoperative day after discharge. Thus, it represents changes in the slope of the outcome trajectory over time.

## References

1. Shichman I, Roof M, Askew N, Nherera L, Rozell JC, Seyler TM, Schwarzkopf R. Projections and Epidemiology of Primary Hip and Knee Arthroplasty in Medicare Patients to 2040-2060. The Journal of bone and joint surgery Open Access 8(1): e22.00112, 2023

2. Bergin PF, Doppelt JD, Kephart CJ, Benke MT, Graeter JH, Holmes AS, Haleem-Smith H, Tuan RS, Unger AS. Comparison of minimally invasive direct anterior versus posterior total hip arthroplasty based on inflammation and muscle damage markers. The Journal of bone and joint surgery American volume 93(15): 1392, 2011

3. Barrett WP, Turner SE, Leopold JP. Prospective randomized study of direct anterior vs postero-lateral approach for total hip arthroplasty. The Journal of arthroplasty 28(9): 1634, 2013

4. Miller LE, Gondusky JS, Bhattacharyya S, Kamath AF, Boettner F, Wright J. Does Surgical Approach Affect Outcomes in Total Hip Arthroplasty Through 90 Days of Follow-Up? A Systematic Review With Meta-Analysis. The Journal of arthroplasty 33(4): 1296, 2018

5. Barrett WP, Turner SE, Murphy JA, Flener JL, Alton TB. Prospective, Randomized Study of Direct Anterior Approach vs Posterolateral Approach Total Hip Arthroplasty: A Concise 5-Year Follow-Up Evaluation. The Journal of arthroplasty 34(6): 1139, 2019

6. Kwon MS, Kuskowski M, Mulhall KJ, Macaulay W, Brown TE, Saleh KJ. Does surgical approach affect total hip arthroplasty dislocation rates? Clin Orthop Relat Res 447: 34, 2006

7. Palan J, Beard DJ, Murray DW, Andrew JG, Nolan J. Which approach for total hip arthroplasty: anterolateral or posterior? Clin Orthop Relat Res 467(2): 473, 2009

8. Maratt JD, Gagnier JJ, Butler PD, Hallstrom BR, Urquhart AG, Roberts KC. No Difference in Dislocation Seen in Anterior Vs Posterior Approach Total Hip Arthroplasty. The Journal of arthroplasty 31(9 Suppl): 127, 2016

9. Booth MW, Riegler V, King JS, Barrack RL, Hannon CP. Patients’ Perceptions of Remote Monitoring and App-Based Rehabilitation Programs: A Comparison of Total Hip and Knee Arthroplasty. The Journal of arthroplasty 38: S58, 2023

10. Sato EH, Stevenson KL, Blackburn BE, Peters CL, Archibeck MJ, Pelt CE, Gililland JM, Anderson LA. Recovery Curves for Patient Reported Outcomes and Physical Function After Total Hip Arthroplasty. The Journal of arthroplasty 38(7s): S65, 2023

11. Eisenach JC, Shields JS, Weller RS, Curry RS, Langfitt MK, Henshaw DS, Pollock DC, Edwards CJ, Houle TT, Spinal Oxytocin Hip Surgery C. Randomized controlled trial of intrathecal oxytocin on speed of recovery after hip arthroplasty. Pain 164(5): 1138, 2023

12. Henshaw DS, Khanna AK, Edwards CJ, Eisenach JC. Hypotension duration and vasopressor requirements following intrathecal oxytocin for Total hip arthroplasty: Secondary analysis of a randomized controlled trial. J Clin Anesth 89: 111189, 2023

13. Shulman SA, Myles PS, Chan MTV, McIlroy DR, Wallace S, Ponsford J. Measurement of Disability-free Survival after Surgery. Anesthesiology 122(3): 524, 2015

14. Higgins BT, Barlow DR, Heagerty NE, Lin TJ. Anterior vs. posterior approach for total hip arthroplasty, a systematic review and meta-analysis. The Journal of arthroplasty 30(3): 419, 2015

15. Nakata K, Nishikawa M, Yamamoto K, Hirota S, Yoshikawa H. A clinical comparative study of the direct anterior with mini-posterior approach: two consecutive series. The Journal of arthroplasty 24(5): 698, 2009

16. Martin CT, Pugely AJ, Gao Y, Clark CR. A comparison of hospital length of stay and short-term morbidity between the anterior and the posterior approaches to total hip arthroplasty. The Journal of arthroplasty 28(5): 849, 2013

17. Zawadsky MW, Paulus MC, Murray PJ, Johansen MA. Early outcome comparison between the direct anterior approach and the mini-incision posterior approach for primary total hip arthroplasty: 150 consecutive cases. The Journal of arthroplasty 29(6): 1256, 2014

18. Christensen CP, Jacobs CA. Comparison of Patient Function during the First Six Weeks after Direct Anterior or Posterior Total Hip Arthroplasty (THA): A Randomized Study. The Journal of arthroplasty 30(9 Suppl): 94, 2015

19. Cheng TE, Wallis JA, Taylor NF, Holden CT, Marks P, Smith CL, Armstrong MS, Singh PJ. A Prospective Randomized Clinical Trial in Total Hip Arthroplasty-Comparing Early Results Between the Direct Anterior Approach and the Posterior Approach. The Journal of arthroplasty 32(3): 883, 2017

20. Boll S, Almeida de Minas AC, Raftogianni A, Herpertz SC, Grinevich V. Oxytocin and Pain Perception: From Animal Models to Human Research. Neuroscience 387: 149, 2018

